# Uncovering Semantic Cognition Beyond Traditional Language batteries: Behavioural and Neural bases of the Semantic Knowledge Test

**DOI:** 10.64898/2025.12.22.25342886

**Authors:** Yoonhye Na, Yu Mi Hwang, JeYoung Jung, Ajay D. Halai, Matthew A. Lambon Ralph, Sung-Bom Pyun

**Affiliations:** Brain Convergence Research Center, Korea University College of Medicine, Seoul, Republic of Korea; School of psychology, University of Nottingham, Nottingham, U.K.; MRC Cognition and Brain Sciences Unit, University of Cambridge, Cambridge, U.K.; Department of Biomedical Sciences, Korea University College of Medicine, Seoul, Republic of Korea; Department of Physical Treatment and Rehabilitation, Korea University Anam Hospital, Seoul, Republic of Korea

**Keywords:** Stroke, Semantic cognition, Aphasia, Assessment, Voxel-based lesion-symptom mapping

## Abstract

Semantic cognition allows us to represent, process and manipulate verbal and nonverbal knowledge about the surrounding environment. Semantic cognition plays a critical role in language, communication and many nonverbal behaviours and can be disrupted by several neurological diseases, including stroke and dementias. Therefore for clinical practice, it is essential to evaluate semantic cognition and to do so using culturally-appropriate assessments (given that some facets of semantic knowledge and associations are culturally-specific). Here, we developed and standardized a new verbal and nonverbal semantic assessment, called semantic knowledge test (SKT), for Korean-speaking populations. The SKT was developed and standardized on a large and demographically diverse sample of 325 healthy adults, confirming its high reliability and validity. To explore its clinical utility and added value, we collected data from 101 post-stroke patients alongside the Western Aphasia Battery (WAB). We run a principal component analysis (PCA) using SKT and WAB scores and Voxel-based lesion-symptom mapping (VLSM) using the factor loadings that yielded from the PCA. The principal component analysis (PCA) of behavioural test scores produced two distinctive factors, one with the WAB scores and the other with the SKT scores. VLSM revealed that the neuroanatomical correlates of the WAB factor covered the peri-Sylvian areas, while that of SKT was in the left superior anterior temporal lobe (ATL) and posterior middle temporal gyrus (pMTG). These findings underscore the utility of the SKT in uncovering both behavioural and neuroanatomical facets of semantic cognition that are not captured by the traditional language batteries such as the WAB and emphasize the importance of independent assessment of semantic deficits in clinical populations.

## Introduction

Stroke is one of the most prevalent cerebrovascular diseases. Cognitive impairment is a frequent symptom across a wide range of neurological diseases including Alzheimer’s disease, semantic dementia, traumatic brain injury, and stroke ^1–3^. Among cognitive functions, semantic cognition allows us to process and manipulate acquired knowledge about the surrounding environment to understand the world and to determine the behaviour ^4^. Semantic cognition is not limited to the language related function but also encompasses non-verbal, multi-modal information including from name, object, face, word and sentence to smell, sound, and texture. Semantic cognition is also disrupted in several neurological diseases, each with distinct neuroanatomical characteristics and different types of difficulties. Semantic cognition plays a critical role in communication and everyday basic functioning. For many decades, semantic assessments for those functional impairments have been developed in Western countries for English-speaking patients. Therefore, it is essential to develop and deploy sensitive and culturally suitable assessments of both verbal and nonverbal semantic cognition for local patients.

Semantic cognition includes two distinctive but interacting parts, semantic memory and semantic control ^4,5^. First, semantic memory is a fundamental component of declarative and explicit memory that encompasses shared knowledge about the world, including words, objects, actions, and concepts ^5,6^. Semantic memory can degrade in disorders such as semantic dementia, resulting from atrophic degeneration of the bilateral anterior temporal lobes (ATL) ^5,7,8^. Semantic control refers to the cognitive processes that manipulate and shape the information associated with each concept ^9,10^. Deficits in semantic control are associated with damage to the left inferior prefrontal cortex or posterior middle and inferior temporal gyri ^9,11^. Thus, stroke-related damage to the frontal and temporoparietal regions can lead to semantic control deficits, where individuals struggle to access and manipulate semantic knowledge despite preserved semantic representations ^5,12,13^.

A variety of semantic cognition assessments have been developed in Western contexts, including well-known tools such as the Pyramids and Palm Trees Test (PPT) and the Camel and Cactus Test (CCT) ^14,15^. These assessments evaluate semantic knowledge using both verbal and nonverbal-picture inputs and have been widely adopted in research and clinical settings ^16–18^. However, the stimuli and conceptual associations are deeply rooted in Western cultural knowledge, which limits their applicability across non-Western populations. Even within Western countries, adaptations and localized norms have been required to maintain validity ^19–22^. In Korea, available tools tend to assess semantic knowledge only indirectly through tasks such as naming, category fluency, or brief language comprehension. These are found in widely used neuropsychological batteries, including the Seoul Neuropsychological Screening Battery (SNSB) ^23^, the Korean Boston Naming Test (K-BNT) ^24^, and the Korean version of the Western Aphasia Battery (K-WAB) ^25^. While these assessments rely on semantic knowledge, they also engage additional processes such as visual perception, language production, and executive function, and thus are not designed to isolate semantic cognition per se. To date, no equivalent tool has been developed for Korean-speaking populations that comprehensively and directly assesses both verbal and nonverbal semantic cognition in a culturally appropriate and theory-driven manner.

To address this gap, the present study developed and validated the Semantic Knowledge Test (SKT), a novel assessment designed to measure verbal and nonverbal semantic cognition in Korean-speaking individuals. Specifically, the study sought to: (i) develop and standardize the SKT; (ii) evaluate its clinical utility in patients with stroke; and (iii) identify the neuroanatomical correlates of language vs. semantic deficits after stroke.

## Method

### Stage I : Development and standardization of the SKT

#### Item selection and scoring

The initial development of the materials to go into the SKT aimed to capture the many diverse semantic relationships between concepts (e.g., synonymous, equivalent, hierarchical (higher/lower), and part-whole relationships). A group of eight experts was organized, comprising linguistics, psycholinguistics, language pathology, and rehabilitation medicine, along with professors in speech therapy, language therapists, and medical doctors specializing in rehabilitation. These experts analyzed existing evaluation tools, previous studies, semantic categories, and semantic associations. Existing tests such as PPT and CCT were analyzed. The PPT includes 52 trials without classified categories, while the CCT comprises 64 trials across four concrete categories. Notably, abstract and part-whole categories are absent in these tools.

Categories relevant to the Korean language were analyzed based on prior studies ^26,27^. From these, 260 target words that represent 20 categories and can be illustrated were extracted. Associations for these words were also investigated. Of the 260 words, 156 were selected based on their potential for pictorial representation. Eight experts rated these items on a three-point scale, and items scoring ≥20 out of 24 points were selected for further refinement. A total of 70 target words and 140 associated words were finalized after excluding synonyms, hierarchical relations, and collocations (Figure 1). These words were carefully chosen to ensure suitability for pictorial representation and cultural relevance. Frequency data for each item were analyzed based on the Sejong Corpus ^28^. The final categories and item distribution are shown in Table 1

**Figure 1.**
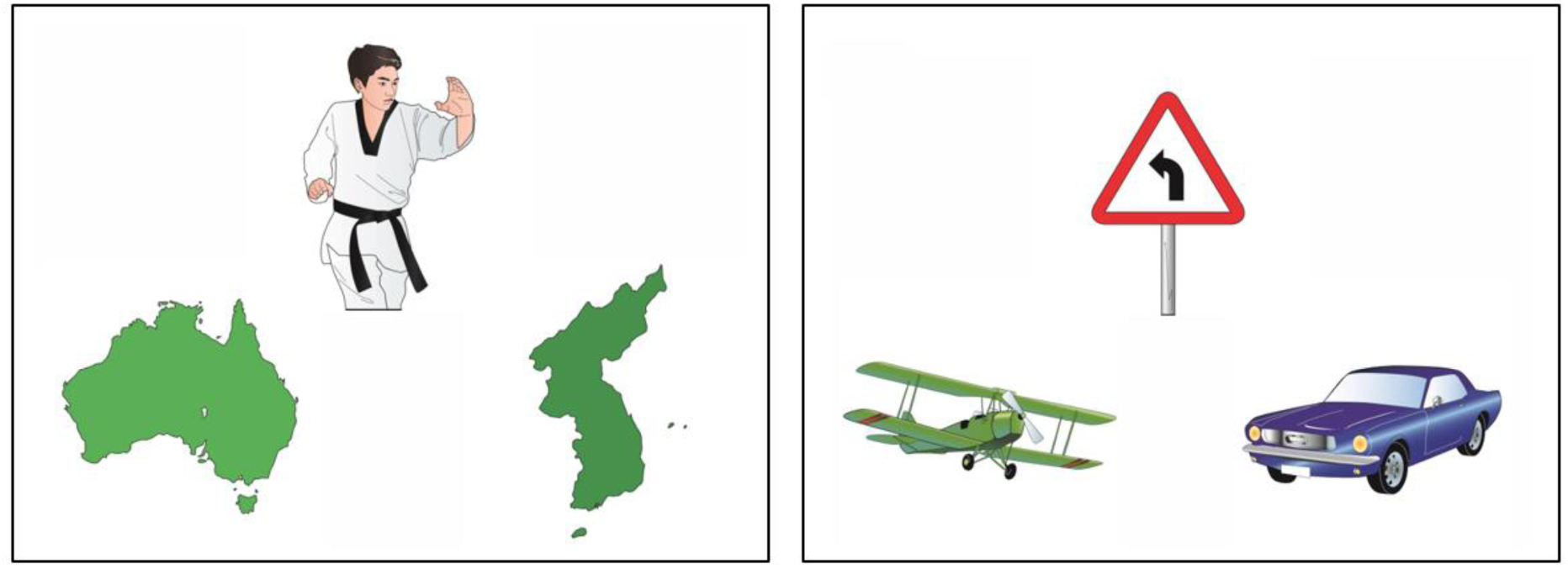
Examples of SKT items (left) culturally biased, (right) culturally universal item

**Table 1.**
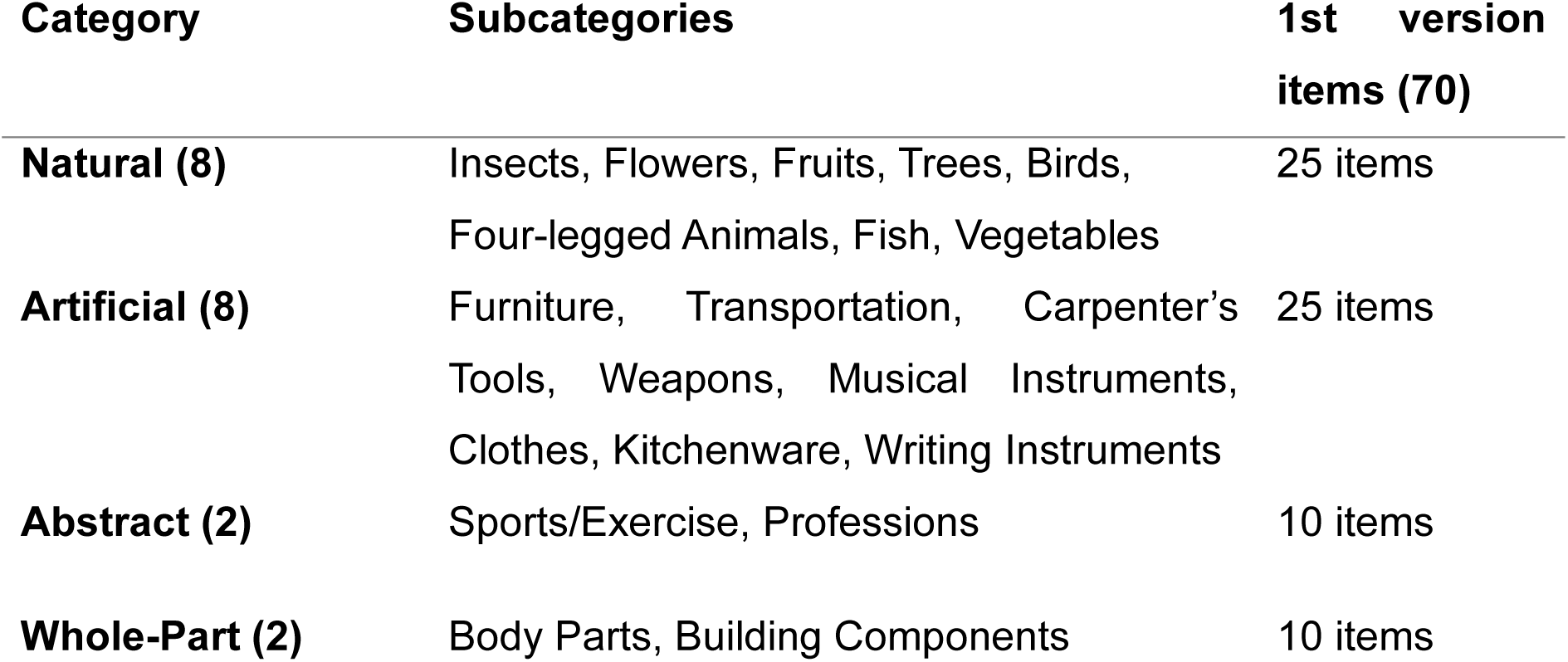
Semantic Categories in the Standardized Test Version (70 Items)

| Category | Subcategories | 1st version items (70) |
| --- | --- | --- |
| **Natural (8)** | Insects, Flowers, Fruits, Trees, Birds, Four-legged Animals, Fish, Vegetables | 25 items |
| **Artificial (8)** | Furniture, Transportation, Carpenter's Tools, Weapons, Musical Instruments, Clothes, Kitchenware, Writing Instruments | 25 items |
| **Abstract (2)** | Sports/Exercise, Professions | 10 items |
| **Whole-Part (2)** | Body Parts, Building Components | 10 items |

**Table 2.** General characteristics of participants (PG1)

| Variables | Healthy controls<br>(N=325) | PG1<br>(N=32) |
| --- | --- | --- |
| Age (yr) | 49.69 ± 18.11 | 56.31 ± 13.47 |
| Gender, male/female | 116 / 209 | 19 / 13 |
| Education (yr) | 13.75 ± 4.08 | 11.13 ± 3.78 |
| K-MMSE (30) | 28.55 ± 1.65 | 20.32 ± 9.16 |
| SKT |  |  |
| Picture test (60) | 58.84 ± 2.35 | 51.50 ± 9.84 |
| Word test (60) | 59.37 ± 1.43 | 56.64 ± 6.15 |
| Pyramids & Palm Trees (52) | - | 36.48 ± 7.72 |

† abbreviations: PG1; patient group 1, K-MMSE; Korean Version of the Mini-Mental State Examination, SKT; semantic knowledge test

A pilot study evaluated 210 images from 70 items. Participants were presented with a target image and two comparison images. They were asked to: (1) select the image more related to the target, and (2) describe the target and comparison images. Based on feedback, the images were revised. To standardize the SKT, data were collected from 325 adults aged 20 and above, along with data from an initial group of 32 brain injury patients for clinical validation.

#### Participants

A total of 325 healthy adults were recruited for inter-item consistency testing and standardization. The inclusion criteria were as follows: (1) age >20 years, (2) a Korean Mini-Mental State Examination (K-MMSE) score within 1 standard deviation of the age norm, and (3) no history of brain injury or neurological or psychiatric disorders. Data from 325 adults were collected nationwide. SKT was administered by trained professionals, including physiatrists, occupational therapists, speech-language pathologists, and psychologists. Data were stratified by age (decades) and education levels for analysis.

To verify the reliability and validity of the SKT, a total of 32 patients (referred to as patient group 1; PG1) with brain damage participated in this study (within 2 months after onset) were recruited. The type of brain damage included stroke (84%), traumatic brain injury (9%), hypoxic brain damage (3%), and dementia (3%). Inclusion criteria were: (1) a confirmed diagnosis via brain imaging, (2) ability to follow SKT instructions, and (3) no prior cognitive impairment. Patients with infratentorial lesions or subarachnoid hemorrhage were excluded.

#### Reliability and validity test

For intra-rater and inter-rater reliability test, two medical professionals independently scored the test, and correlation analysis was conducted. For test-retest reliability, the test was re-administered after two weeks, and the correlation between scores was analyzed. For validity testing, concurrent validity was assessed by correlating SKT scores with PPT scores.

#### Statistical analysis

Data analysis was performed using SPSS Statistics version 26 (IBM, Armonk, NY, USA). Statistical significance was set at P < 0.05. Internal consistency was assessed using Cronbach’s alpha. Pearson’s correlation was used for relationships between SKT scores, age, and education. Spearman’s correlation was used for intra- and inter-rater reliability. Post-hoc analysis of SKT scores was conducted by age and education groups. Concurrent validity was measured by correlating K-MMSE and PPT scores.

### Stage II: Identifying clinical utility of SKT and neuroanatomical correlates in stroke

#### Stroke patient enrollment

We enrolled stroke patients (referred as patient group 2; PG2) retrospectively from the STroke Outcome Prediction database (STOP DB; Department of Physical Treatment and Rehabilitation, Korea University Anam Hospital, Seoul, Republic of Korea) which is registered on the Clinical Research information Service (CRIS; registration number : KCT0006047). We reviewed the STOP DB from November 2014 to November 2023. The criteria for inclusion were as follows; i) aged between 20 and 85, ii) who completed the SKT and K-WAB with both versions of picture and word within 6 months after the infarction, iii) MRI scanning was undergone within 6 months after onset, iv) patients with the first-ever unilateral stroke (left or right) confirmed by brain imaging (e.g., MRI or CT). The exclusion criteria were as follows; i) had brain damage other than stroke including traumatic brain injury, or brain tumor, ii) had the lesions in the infratentorial regions, iii) had a prior cognitive, neurologic or psychiatric medical history, iv) showed a cognitive decline before the onset of the stroke.

A total of 101 patients with unilateral stroke were included in the study. We collected patients’ behavioral test results, MR images and demographic data including age, sex, years of education, handedness and time post onset from the STOP DB. Table 3 describes the demographic and behavioral characteristics of PG2. The mean age of the subjects was 60.19 (SD = 14.25), and MRI scanning was conducted 31.18 days on average after stroke onset. 67 patients had lesions in the left hemisphere and the others had lesions in the right hemisphere.

**Table 3.** Demographic and behavioral characteristics of PG2 (N=101)

|  | Mean (SD) | Range |
| --- | --- | --- |
| Age (years) | 60.19 (14.25) | 21 - 86 |
| Education (years) | 12.78 (4.01) | 3 - 21 |
| Time post onset (days) | 31.18 (20.89) | 2 - 101 |
| Lesion volume (mm<sup>3</sup>) | 51293.77<br>(60789.48) | 1,550 – 338,200 |
| Lesion side (L/R) | 67 / 34 |  |
| Sex (M/F) | 58 / 43 |  |
| SKT |  |  |
| SKT-p | 51.94 (11.57) | 0 - 60 |
| SKT-w | 51.26 (12.86) | 0 - 60 |
| K-MMSE | 19.48 (8.44) | 1 - 30 |
| K-WAB (AQ) | 68.54 (29.42) | 0.20 – 99.20 |
| Speech fluency | 13.89 (5.92) | 0.00 – 20.00 |
| Functional content | 6.77 (3.17) | 0.00 – 10.00 |
| Fluency | 7.12 (2.97) | 0.00 – 10.00 |
| Comprehension | 7.05 (2.90) | 0.00 – 10.00 |
| yes-no | 48.06 (16.40) | 0.00 – 60.00 |
| Auditory word recognition | 45.65 (17.94) | 0.00 – 60.00 |
| Sequential command | 47.29 (28.06) | 0.00 – 80.00 |
| Repetition | 6.98 (3.56) | 0.00 – 10.00 |
| Repetition | 68.96 (36.05) | 0.00 – 100.00 |
| Naming | 6.37 (3.46) | 0.00 – 10.00 |
| Object naming | 42.70 (22.00) | 0.00 – 60.00 |
| Word fluency | 7.30 (6.18) | 0.00 – 20.00 |
| Sentence completion | 6.96 (4.04) | 0.00 – 10.00 |
| Responsive speech | 6.73 (4.19) | 0.00 – 10.00 |

\* PG2; patient group 2, SKT; semantic knowledge test for adults, K-MMSE; Korean version of mini-mental state examination, K-WAB; Korean version of Western aphasia battery, AQ; aphasia quotient

#### Principal component analysis (PCA)

In this study, principal components were extracted from the patients’ test results from the SKT and K-WAB. All scores from the SKT-p, SKT-w, and each subtest of the K-WAB were entered into a PCA with varimax rotation using IBM SPSS Statistics (Version 25.0). The two factors with an eigenvalue >1 were extracted. The patients’ factor scores on these two principal components were used in the lesion-symptom mapping analyses.

#### Image acquisition

Every patient underwent MRI scanning within 6-month after the onset of stroke. We collected the high-resolution structural T1-weighted MRI, B0 image from the diffusion tensor image (DTI). All MR images were acquired on a 3.0T Siemens Prisma scanner (Siemens, Erlangen, Germany) at Korea University Anam Hospital.

A T1-weighted image was acquired with the following parameters: repetition time (TR) = 2020 ms, echo time (TE) = 2.91 ms, flip angle = 9°, filed of view (FOV) = 217 × 166 mm^2^, voxel size = 0.34 × 0.8 × 0.34 mm^3^. The DTI was acquired with 64 diffusion gradient directions and the following acquisition parameters: repetition time (TR) = 6500 ms, echo time (TE) = 55 ms, flip angle = 90°, field of view (FOV) = 224 x 224 mm^2^, voxel size = 1.0 x 1.0 x 2.0 mm^3^, b = 1000 sec/mm^2^.

#### VLSM analysis

We manually identified the lesion of stroke from each MR image of patients using ITK-SNAP ver. 3.8.0. ^29^. The expert researchers (YN and MC) performed manual lesion identification on the B0 image, the first volume of the diffusion tensor image (DTI), with visual reference to the FLAIR image. Lesion binary images were coregistered with the individual T1 image using the SPM12 toolbox (https://www.fil.ion.ucl.ac.uk/spm/software/spm12) implemented in MATLAB (R2019a, The MathWorks, Inc., Natick, Massachusetts, United States), and then the coregistered images were normalized to the standard MNI152 brain. We also measured the lesion volume of each patient from the lesion binary image and used it as a confounding variable of no interest for the statistical model. The lesion overlay map was built by summing all the individual lesion binary maps using SPM12. One case was excluded due to the failure in the spatial normalization process, thus, 100 cases were included in the lesion-symptom mapping.

Normalized lesion binary images were entered into the statistical modeling. Since the lesions and behavioral data of patients did not follow the normal distribution, the statistical analysis was conducted using a Statistical NonParametric Mapping (SnPM; version 13.1.08, http://nisox.org/Software/SnPM13/) toolbox implemented in MATLAB to run the non-parametric analysis. We entered several confounding factors including age, sex, year of education and lesion volume into the statistical modeling. The whole-brain analysis results were considered significant at P_FWE_ < .05 after multiple comparison correction.

Regions of interest (ROI) were selected from previously known areas associated with semantic processing including the bilateral IFG, pMTG, and ATL (Figure 2) ^5,7–9,11–13^. ROI masks of the IFG, pMTG and ATL were selected from anatomically predefined atlases using the WFU_PickAtlas toolbox (version 3.0.5, https://www.nitrc.org/projects/wfu_pickatlas) ^30,31^. ROI mask for the pMTG was revised and used from the whole MTG map. Selected ROI masks were used as an explicit mask when specifying each statistical model. All the results were visualized and reported using MRIcroGL (version 1.2.20211006, https://www.nitrc.org/plugins/mwiki/index.php/mricrogl).

**Figure 2.**
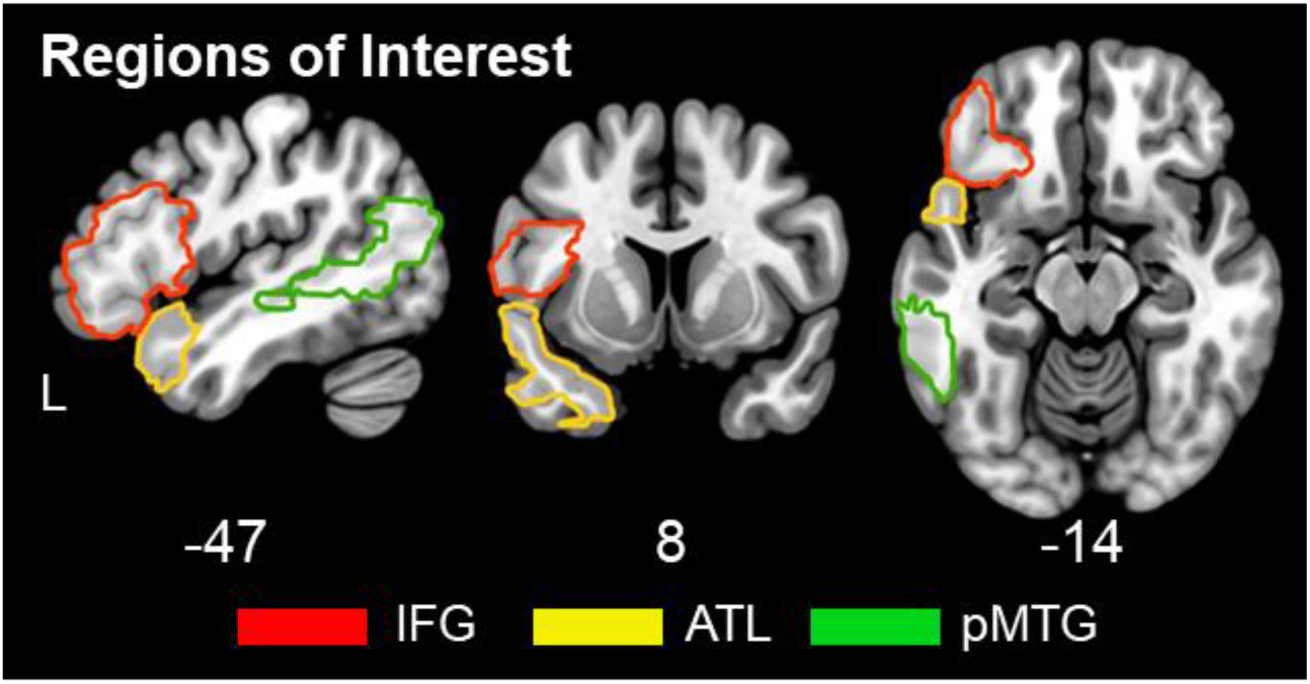
The location of ROIs. (The identical regions to the ROIs in the left hemisphere were used to place ROIs in the right hemisphere)

#### Lesion overlay map

A lesion overlay map for patients with stroke is provided in Figure 3. Most of the patients’ lesions fell into the territory supplied by the middle cerebral artery (MCA). A minority of patients had lesions in the territory of the anterior or posterior cerebral arteries (ACA and PCA, respectively), and some overlaid regions were extended in the areas affected by two arteries. There were more overlapping affected regions in the left hemisphere since there were double the number of left-side damaged patients (66.3%) than right (33.7%).

**Figure 3.**
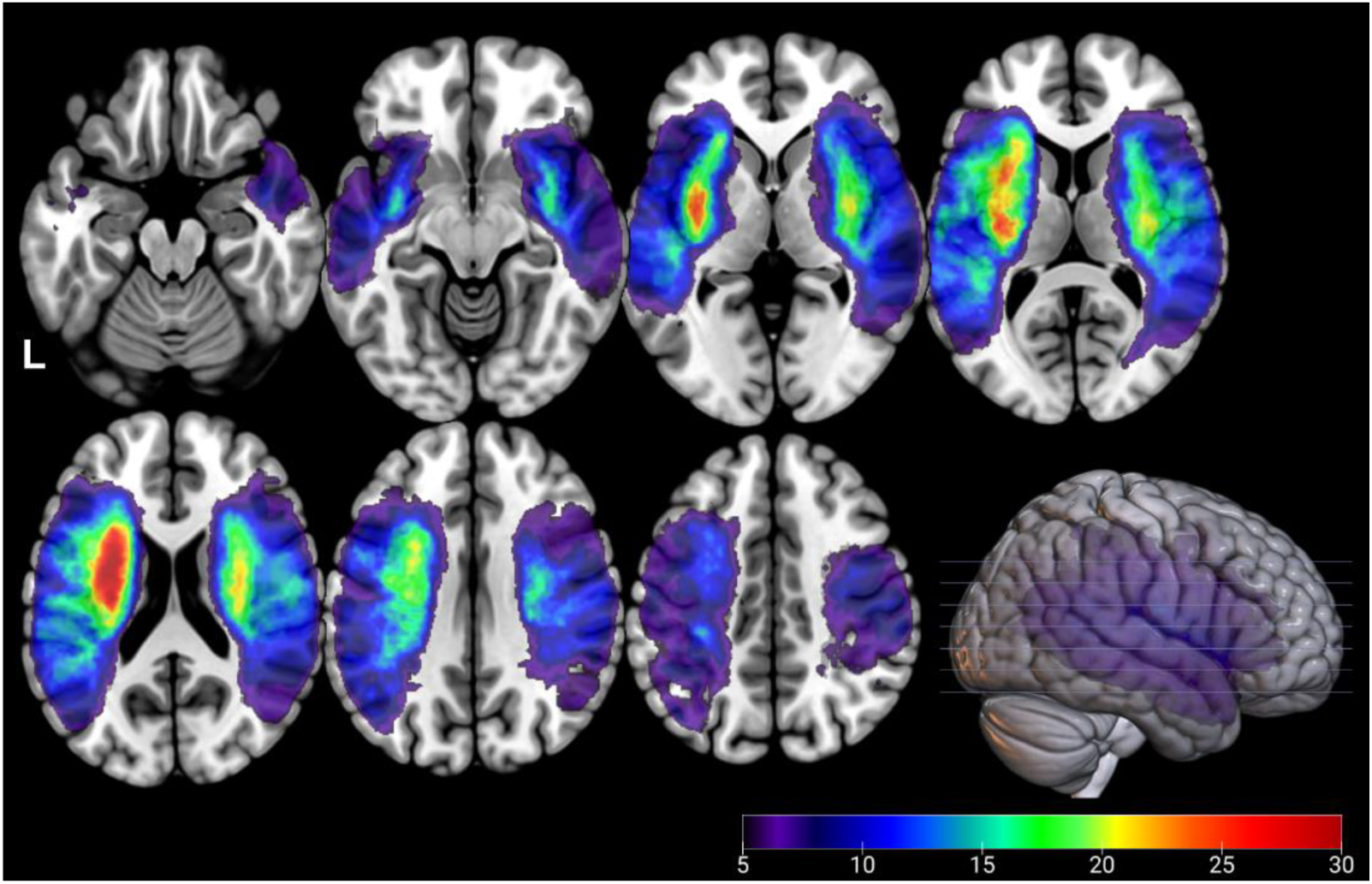
Lesion overlay map of 101 patients with stroke. The color bar placed in the right bottom indicates how many lesions overlapped.

## Results

### Stage I : Development and standardization of the SKT

A total of 325 healthy participants (116 males, 209 females) were enrolled for the standardization of the SKT. The mean age of the participants was 49.69 years, with a mean education duration of 13.75 years. The mean K-MMSE score was 28.55 points.

For the semantic knowledge test, 60 final items were selected (Appendix 1). Internal consistency (Cronbach’s alpha) of the initial 70-item test version was analyzed, yielding a value of 0.792 for the picture test and 0.674 for the word test. Items with ambiguous or difficult-to-judge images (e.g., pumpkin-carriage, window-curtain) were excluded through expert consensus. Subsequently, items that improved internal consistency when removed were sequentially eliminated. This process resulted in a final set of 60 items, with internal consistency values improving to 0.815 for the image test and 0.721 for the word test.

The mean scores for the picture test and word test were 58.84 (out of 60) and 59.37 (out of 60), respectively. Pearson correlation analysis revealed significant relationships between SKT performance and participant age and education level. For the picture test, performance was negatively correlated with age (r = −0.499, P < 0.001) and positively correlated with education level (r = 0.521, P < 0.001). Similarly, for the word test, performance was negatively correlated with age (r = −0.465, P < 0.001) and positively correlated with education level (r = 0.519, P < 0.001). Table 4 shows performance summaries on the test split by education and age.

**Table 4.** SKT test scores according to age and education groups (N=325)

|  | N | SKT-p (60) | SKT-w (60) |
| --- | --- | --- | --- |
| *Age group* |  |  |  |
| 20 - 29 | 63 | 59.71 ± 0.79 | 59.84 ± 0.37 |
| 30 - 39 | 58 | 59.86 ± 0.40 | 59.95 ± 0.22 |
| 40 - 49 | 38 | 59.55 ± 1.48 | 59.74 ± 1.16 |
| 50 - 59 | 62 | 59.21 ± 1.33 | 59.65 ± 0.72 |
| 60 - 69 | 43 | 58.86 ± 1.52 | 59.23 ± 1.02 |
| > 70 | 61 | 56.13 ± 3.82 | 57.92 ± 2.44 |
| *Education group* |  |  |  |
| 0 | 2 | 52.50 ± 3.54 | 56.00 ± 0.00 |
| 1-6 | 32 | 55.28 ± 4.24 | 57.22 ± 2.88 |
| 7-9 | 24 | 58.04 ± 1.94 | 58.83 ± 1.31 |
| 10-12 | 75 | 58.81 ± 2.12 | 59.35 ± 1.25 |
| ≥13 | 192 | 59.61 ± 1.00 | 59.84 ± 0.43 |

### Reliability and validity of the SKT

#### Reliability and validity

Inter-rater reliability was extremely high, with correlation coefficients of *r* = 0.972 for the picture test and *r* = 1.0 for the word test (*P* < 0.001). Intra-rater reliability was also high, with *r* = 0.838 for the picture test and *r* = 0.987 for the word test (*P* < 0.001).

Concurrent validity was demonstrated through the correlation of SKT scores with PPT scores in PG1, which produced a correlation coefficient of *r* = 0.902 for SKT-p (*P* = 0.001) (Figure 4).

**Figure 4.**
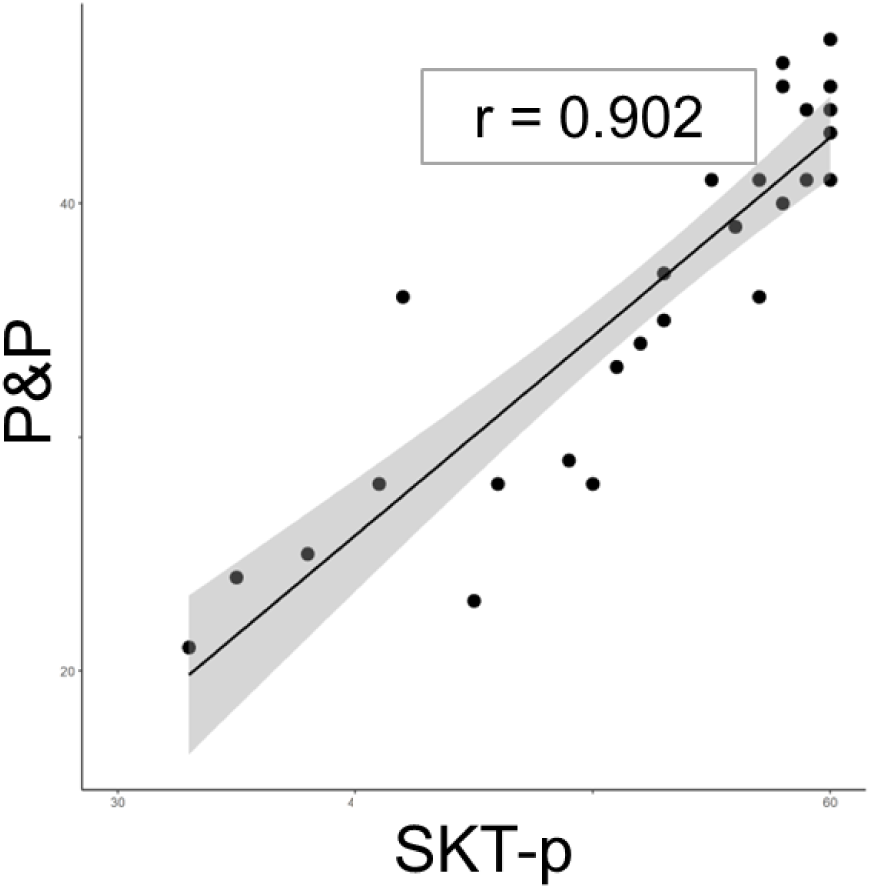
Scatter plots between SKT-p and P&P

### Stage II: Identifying clinical utility of SKT and neuroanatomical correlates in stroke

#### PCA results

Two independent principal factors were yielded from the PCA, and the two factors accounted for 80.88% of variance in performance of patients (factor 1 = 69.73%, factor 2 = 11.15%). The Kaiser–Meyer–Olkin measure of sampling adequacy was 0.904. Each factor loading is shown in Table 5. The PCA result showed that all the subtest performances from each K-WAB subtest loaded heavily on Factor 1 (thus referred to as “Factor_WAB”). The two SKT scores combined into Factor 2 (thus named “Factor_SKT”). This PCA result indicated that the two assessments successfully probed the different language and semantic domains, rather than picking up on one single severity based variation. Scores on Factor 1 were highly correlated with WAB AQ (r = 0.961, p<.001), indicating that this factor is a data-driven aphasia severity measure.

**Table 5.** Factor loadings.

|  | Factor 1<br>(WAB) | Factor 2<br>(SKT) |
| --- | --- | --- |
| Object naming | .917 | .176 |
| Functional content | **.907** | .235 |
| Responsive speech | **.906** | .254 |
| Sentence completion | **.900** | .278 |
| Fluency | **.853** | .058 |
| Repetition | **.838** | .287 |
| Word fluency | **.800** | .201 |
| Sequential command | **.785** | .325 |
| Auditory word recognition | **.759** | .428 |
| Yes-No question | **.739** | .366 |
| SKT-p | .134 | **.954** |
| SKT-w | .339 | **.885** |

#### VLSM results

##### Whole-brain analysis

Figure 5 and Table 6 show the results of the VLSM using whole-brain analysis. All the VLSM results were significant only in the left hemisphere.

**Figure 5.**
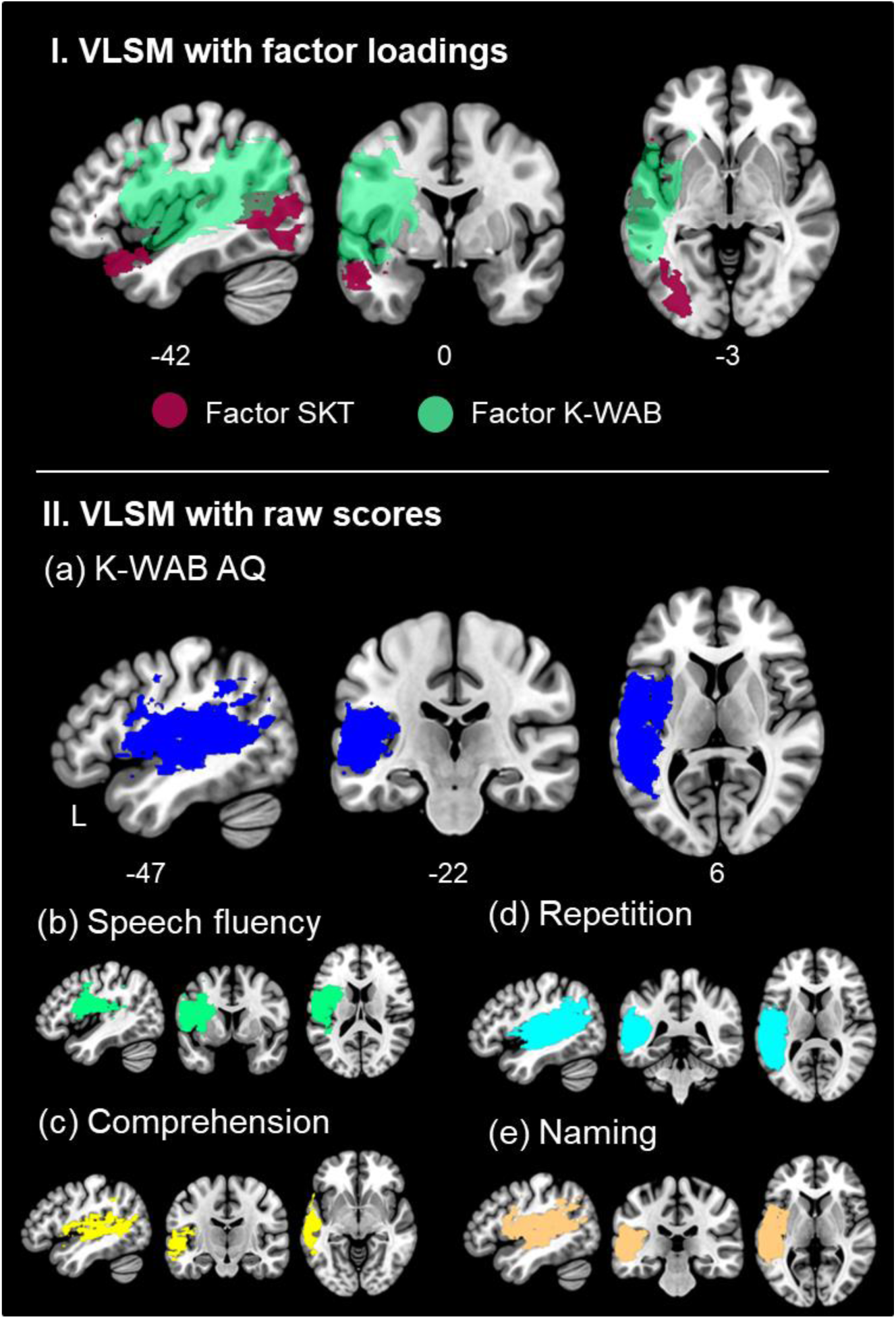
VLSM results with AQs from K-WAB (I) (a) K-WAB AQ, (b) speech fluency, (c) comprehension, (d) repetition, (d) naming and VLSM result with factor scores (II)

**Table 6.** The VLSM results with PCA factors and AQs of K-WAB.

| Measure | Cluster<br>size | T | $P_{FWE}$ | Coordinates | | | Label |
| --- | --- | --- | --- | --- | --- | --- | --- |
|  |  |  |  | x | y | z |  |
| *PCA factors* |  |  |  |  |  |  |  |
| Factor | 8926 | 6.76 | 0.0918 | -66 | -12 | 2 | Lt. middle temporal gyrus |
| **SKT** |  |  |  | -58 | -62 | 8 |  |
|  |  |  |  | -40 | -60 | 0 | Lt. middle occipital gyrus |
| **Factor** | 6661 | 7.81 | 0.0002 | -48 | -14 | 4 | Lt. superior temporal |
| **K-WAB** |  |  |  | -58 | -12 | 12 | gyrus |
|  |  |  |  | -48 | -32 | 14 |  |
| **K-WAB scores** |  |  |  |  |  |  |  |
| **K-WAB** | 6719 | 8.06 | 0.0002 | -44 | -32 | 14 | L. superior temporal gyrus |
| **AQ** |  |  |  | -58 | -12 | 12 |  |
|  |  |  |  | -48 | -14 | 4 | L. Heschl's gyrus |
| **Speech** | 3022 | 8.32 | 0.0002 | -32 | 2 | 16 | L. insula |
| **fluency** |  |  |  | -36 | -6 | 16 |  |
|  |  |  |  | -52 | 4 | 32 | L. precentral gyrus |
| **Compreh** | 2938 | 7.29 | 0.0002 | -56 | 8 | -2 | L. temporal pole |
| **ension** |  |  |  | -44 | -34 | 14 | L. superior temporal gyrus |
|  |  |  |  | -64 | -10 | -2 |  |
| **Repetitio** | 8931 | 11.10 | 0.0002 | -46 | -46 | 16 | L. middle temporal gyrus |
| **n** |  |  |  | -48 | -32 | 14 | L. superior temporal gyrus |
|  |  |  |  | -60 | -20 | 10 |  |
| **Naming** | 6742 | 8.03 | 0.0002 | -46 | -46 | 16 | L. middle temporal gyrus |
|  |  |  |  | -48 | -28 | 4 | L. superior temporal gyrus |
|  |  |  |  | -58 | -12 | 4 |  |

The VLSM results for the PCA factors revealed distinct patterns between the two factors. The patients’ Factor SKT scores were related to clusters included the left anterior superior ATL and the posterior lateral temporal region. The cluster associated with Factor K-WAB scores covered the core perisylvian region (Figure 5-I). As expected given their high correlation, the result for the Factor_K-WAB and the K-WAB AQ scores was almost identical (Figure 5-II). Although the different subtests of the K-WAB all correlated highly with a single principal component, we conducted additional exploratory analyses for each language task. The results are shown in Figure 5-II. Consistent with the high correlations, performance on these language probes all loaded into the same perisylvian territory, with some subtle (but not significant) differences in extent.

##### ROI analysis

In line with the whole-brain results, the ROI analyses found that tissue integrity in both the left ATL and MTG were significantly correlated with SKT performance. Scores on Factor K-WAB showed the significant relationship with all the ROIs in the left hemisphere including IFG, ATL and MTG (Table 7).

**Table 7.**
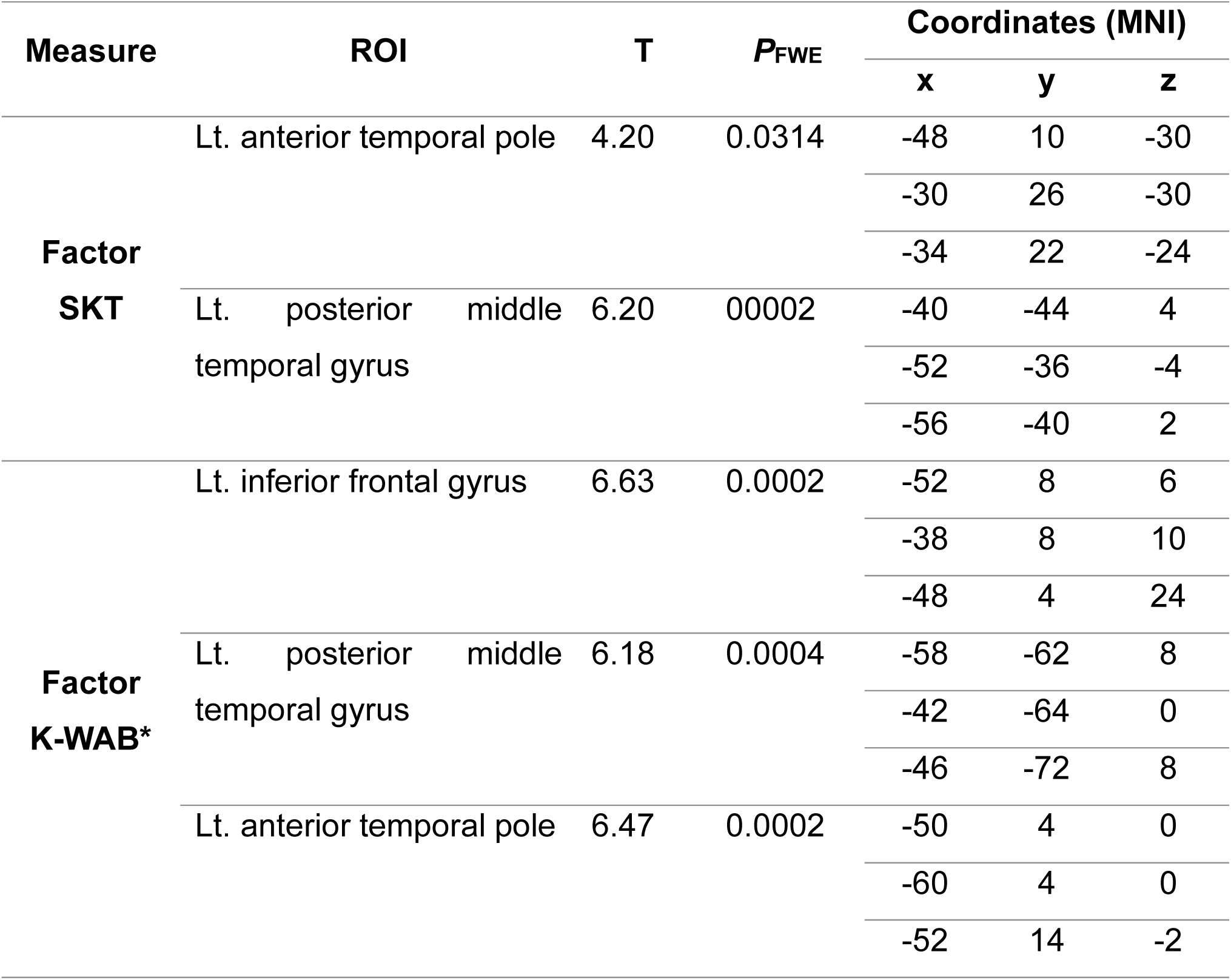
The results of ROI analysis.

## Discussion

We developed and validated a new semantic test, the Semantic Knowledge Test (SKT), as a standardized assessment tool to evaluate semantic cognition both verbally and nonverbally. We then applied the SKT to a stroke patient cohort to assess its clinical utility in comparison to the widely used Western Aphasia Battery (WAB), and to identify the neuroanatomical correlates of post-stroke semantic and language deficits. Our findings suggest that the SKT captures a distinct aspect of comprehension—specifically semantic cognition—that is not effectively detected by the traditional language batteries such as the WAB. Furthermore, lesion mapping revealed that the neuroanatomical patterns associated with performance on the SKT differed from those associated with the WAB. This indicates that the two tests, as planned, tap into different cognitive domains that are mediated by distinct neural substrates.

The SKT was developed as a novel, psycholinguistically informed assessment to address the lack of culturally appropriate tools for the assessment of semantic cognition in Korean-speaking populations. Standardized on a large and demographically diverse sample of 325 healthy adults, the SKT demonstrated strong psychometric properties, including high internal consistency and excellent inter- and intra-rater reliability. Its application to an initial group of 32 patients with brain injury confirmed its clinical sensitivity, as these patients scored significantly lower than healthy controls. Moreover, the SKT showed strong concurrent validity, with a high correlation (r = .90) with the PPT, and it successfully captured semantic impairments not detected by general language assessments such as the WAB. These findings support the SKT as a theoretically grounded and clinically valuable tool that reveals subtle semantic deficits, which may otherwise remain unrecognized in standard assessments.

The PCA of the data collected from the large N=100 patient cohort showed that the SKT and WAB are underpinned by two independent sources of variation. The high loadings of all K-WAB subtests on Factor 1 and its very high correlation with WAB AQ indicate that this component directly reflect aphasia severity. Replicating other recent results, the second independent factor spanned both the verbal and nonverbal version of the SKT without loadings from the K-WAB language subtests, indicating that performance on multimodal semantics dissociates away from other aspects of language and other cognitive functions ^32–34^. The VLSM results showed that the clusters correlated with K-WAB, the K-WAB AQ and its sub-test scores covered the same broad perisylvian region, with no clear differences between the four different K-WAB sub-tests. This result would seem to stem from the fact that, although probing different language tasks, performance in these subtests across the 100 patients is very highly correlated and thus primarily reflects aphasia severity alone. Taking the behavioural results and lesion correlates together, this study replicates previous investigations which indicated that broad language screeners tend to be intensive to the presence and severity of semantic impairment, but that these deficits can be identified and quantified through targeted semantic tests or batteries such as the Cambridge Semantic Battery (CSB) ^5,35^.

The VLSM for the semantic factor identified both the left superior ATL and posterior lateral temporal lobe (extending from middle temporal to lateral temporo-occipital gyrus). This posterior lateral region overlaps with the posterior aspects of the semantic control network as highlighted in studies of semantic aphasia ^5^, as well as rTMS and fMRI in healthy participants ^36–39^. The identified superior ATL area does not overlap precisely with ATL regions implicated in semantic representation, which tend to be centred on the more ventrolateral aspects of this region by fMRI and studies of semantic dementia ^40^. This is likely to reflect the nature of the vascular supplies: the middle cerebral artery (MCA) does not perfuse the mid to ventral ATL and thus these regions are rarely damaged after embolic stroke ^41^.

## Conclusion

In conclusion, this study developed a new culturally-appropriate semantic knowledge test to evaluate verbal and nonverbal semantic cognition. Using this test alongside the K-WAB in a large group of post-stroke aphasic patients, allowed for the identification of both aphasia severity and, independently, multimodal semantic performance. Our findings highlight the necessity of evaluating semantic deficits separately, particularly in clinical populations where deficits in lexical-semantic processing may not be adequately detected by WAB’s comprehension measures.

## Data availability

The data used in this study are available upon reasonable request from the corresponding authors. The data are not publicly available as they include information that may compromise participant confidentiality.

## Research ethics

This study protocol was approved by the appropriate institutional review board of Korea University Anam Hospital (IRB no. 2014AN14145, 2019AN0293) and conducted in accordance with the principles of the Declaration of Helsinki.

## Patient consent

This study was conducted with the approval by the Institutional Review Board (IRB) committee at Korean University Anam Hospital (IRB no. 2025AN0050). As this study was conducted restrospectively, the requirement for informed consent from patients was waived.

